# Algorithmic Identification of Potentially High Risk Abdominal Presentations (PHRAPs) to the Emergency Department: A Clinically-Oriented Machine Learning Approach

**DOI:** 10.1101/2022.02.08.22270691

**Authors:** Richard S Kuzma, Varun Saraswathula, Kathryn R Moon, Rachel R Kelz, Ari B Friedman

## Abstract

**Background:** Older adults presenting to emergency departments (EDs) with abdominal pain have been shown to be at high risk of subsequent morbidity and mortality. Yet, such presentations are poorly studied in national databases. Claims databases do not record the patient’s symptoms at the time of presentation to the ED, but rather the diagnosis after testing and evaluation, limiting study of care and outcomes for these high risk abdominal presentations.

**Objectives:** We sought to develop an algorithm to define a patient population with potentially high risk abdominal presentations (PHRAPs) using only variables commonly available in claims data.

**Research Design:** Train a machine learning model to predict abdominal pain chief complaints using the National Hospital Ambulatory Medical Care Survey (NHAMCS), a nationally-representative database of abstracted ED medical records.

**Subjects:** All patients contained in NHAMCS data from 2013-2018. 2013-2017 were used for predictive modeling and 2018 was used as a hold-out test set.

**Measures:** Positive predictive value and sensitivity of the predictive algorithm against a hold-out test set of NHAMCS patients the algorithm was blinded to during training. Predictions were assessed for agreement with either a chief complaint of abdominal pain (contained in “Reason for Visit 1”), or an expanded definition intended to capture visits which were for abdominal concerns. These included secondary or tertiary complaints of abdominal pain or other abdominal conditions, other abdominal-related chief complaint (e.g. nausea or diarrhea, but not pain), discharge diagnosis of an abdominal condition, or reception of an abdominal CT or ultrasound.

**Results:** After validation on a hold-out data set, a gradient boosting machine (GBM) was the best best-performing machine learning model, but a logistic regression model had similar performance and may be more explainable and useful to future researchers. The GBM predicted a chief complaint of abdominal pain with a positive predictive value of 0.60 (95% CI of 0.56, 0.64) and a sensitivity of 0.29 (95% CI of (0.27, 0.32). Nearly all false positives still exhibited signs of “abdominal concerns” for patients: using the expanded definition of “abdominal concern” the model had a PPV of >0.99 (95% CI of 0.99, 1.00) and sensitivity of 0.12 (95% CI of 0.11, 0.13).

**Conclusion:** The algorithm we report defines a patient population with abdominal concerns for further study of treatment and outcomes to inform the development of clinical pathways.

## Introduction

### Background

Older adults with chief complaints of abdominal pain presenting to emergency departments (EDs) experience mortality as high or higher than ST-elevated myocardial infarction (STEMI).^1,2^ Despite the high risks of chief complaints of abdominal pain in these patients, 40% do not get a CT nationally.^3^ By comparison, 93% of 30-39 year olds with a chief complaint of chest pain receive an EKG (authors’ calculations). Unlike for chest pain, no acute activation pathway exists to prioritize and systematize care for chief complaints of abdominal pain.

Conceptually, a chief complaint in the ED captures a syndromic presentation based on the patient’s own understanding of their symptoms and disease process; it has been used to define research and clinical protocols, similar to how consensus definitions of disease (e.g. fibromyalgia) were instrumental in understanding and improving care.^4^ A chief complaint is symptom-driven and patient-centered rather than disease- or biology-centric, and is of central importance in studying the quality and nature of emergency department care.^4^ Unfortunately, no single data source exists that shows post-discharge outcomes for the patient population that entered an emergency department with a chief complaint of abdominal pain. National Hospital Ambulatory Medical Care Survey (NHAMCS) data includes chief complaints, diagnostic tests, and diagnoses; Medicare data includes post-discharge outcomes and diagnostic tests but not chief complaints. Previous research indicates value in defining patient populations for clinical and health services research using administrative datasets.^5^ Better understanding the outcomes of patients with a chief complaint of abdominal pain will allow researchers to better quantify the risks, define metrics for success, and define care pathways and quality metrics for these high-risk cases.

Machine learning techniques have become established tools in clinical and health services research because they allow non-linear pattern finding without overfitting.^6–9^ We apply machine learning techniques to abdominal presentations to the ED.

### Intended Audience

We propose a novel algorithm to determine whether an ED visit was for a potentially high-risk abdominal presentation (PHRAP). The algorithm is intended for research, rather than clinical purposes. To do so, we first trained an algorithm on NHAMCS data to predict a chief complaint of abdominal pain and validate the model on a hold-out test set of unseen NHAMCS data. Future use of our algorithm on Medicare data will help researchers define and study a patient population with abdominal symptoms at high risk of subsequent complications that was previously unavailable to study outcomes and risk, and how care patterns modify that risk.

In developing the model, we optimized model parameters and methods that help to increase positive predictive value at the expense of sensitivity. This will likely subset for more clearly abdominal and potentially higher-risk presentations; determining how the population defined by PHRAP correlates with a chief complaint of abdominal pain and with risk of post-discharge mortality will require further research.

We also conduct error analysis of our PHRAP model’s false positive predictions to see whether the model is still picking up on abdominal complaints. This way, future researchers may understand that false positive predictions are still relevant when tracking outcomes of abdominal complaints in claims data.

## Data

The study population consists of NHAMCS data of adults 18 years and older who visited U.S. EDs from 2013-2018 (n=96,593). We included only variables available in Medicare claims data (where the algorithm is intended for use). We used the “Reason for Visit 1” variable to define a binary classification problem. The patient was labeled as a “Chief Complaint of Abdominal Pain” if any of the following abdominal pain complaints in Table 1 occurred within “Reason for Visit 1”:

- Abdominal Pain cramps, spams, not otherwise specified
- Lower abdominal pain, cramps, spasms, not otherwise specified
- Upper abdominal pain, cramps, spasms, not otherwise specified
- Stomach and abdominal pain, cramps, spasms

**Table 1.** Definitions of Chief Complaint of Abdominal Pain and Abdominal Concern.

| Chief Complaint of Abdominal Pain | Abdominal Concern |
| --- | --- |
| <u>RFV<sub>1</sub> contains any of:</u> | <u>RFV<sub>1</sub> or RFV<sub>2</sub> contains any of:</u> |
| Abdominal Pain cramps, spasms, NOS<br>Lower abdominal pain, cramps, spasms, NOS<br>Upper abdominal pain, cramps, spasms, NOS<br>Stomach and abdominal pain, cramps, spasms | Abdominal pain, cramps, spasms, NOS<br>Lower abdominal pain, cramps, spasms, NOS<br>Upper abdominal pain, cramps, spasms, NOS<br>Stomach and abdominal pain, cramps, spasms<br><u>RFV<sub>1</sub> or RFV<sub>2</sub> contains any of:</u><br>Hernia of abdominal cavity<br>Abdominal distension, fullness, NOS<br>Bladder pain, Nausea, Vomiting, Diarrhea<br>Constipation, Abdominal size change, Abnormal appetite,<br>Pain of liver, gallbladder, and biliary system<br><u>Discharge Diagnosis:</u><br>Discharge diagnoses found in chief complaint of abdominal pain<sup>3</sup> then curated based on clinical judgment to only abdominal diagnoses (Appendix Table 2)<br><u>Imaging to include:</u><br>CT of abdomen<br>Ultrasound of abdomen |

## Methods

We trained our machine learning model to make PHRAP predictions using 2013-2017 NHAMCS data (N=81,008)—allowing our model to “learn” from data, make a classification prediction of “PHRAP” or “non-PHRAP”, and receive feedback from patients actually labeled as a true case of PHRAP or not.

Descriptive statistics of the training and testing set are found in Table 2, in the results section.

**Table 2.** Population description for training and test data sets. Source: NHAMCS, 2013-2018.

| **Variable** | **Training Set**<br>2013-2017<br>n=81,008, % | **Test Set**<br>2018<br>n=15,585, % | **P-value** |
| --- | --- | --- | --- |
| Age (years), (SD) | 46.5<br>(19.66) | 47.7<br>(19.86) | <0.001 |
| **Ethnicity** |  |  |  |
| Hispanic/Latino | 10.3 | 10.9 | 0.02 |
| Not Hispanic/Latino | 65.25 | 69.25 | <0.001 |
| Blank | 24.5 | 19.9 | <0.001 |
| **Race** |  |  |  |
| White | 60.6 | 56.75 | <0.001 |
| Black | 19.8 | 21.4 | <0.001 |
| Blank | 16.7 | 18.6 | <0.001 |
| Asian | 1.72 | 1.73 | 0.93 |
| Other | 1.20 | 1.46 | <0.001 |
| **Female %** | **43.01** | **43.68** | 0.12 |
| **Payer type, %** |  |  |  |
| Medicare | 23.3 | 24.04 | .046 |
| Private Insurance | 36.5 | 33.33 | <0.001 |
| Unspecified/Other | 12.15 | 14.29 | <0.001 |
| Medicaid | 29.7 | 31.89 | <0.001 |
| Self-pay | 13.4 | 11.11 | <0.001 |
| No charge/charity | 1.19 | 0.82 | <0.001 |
| No Payment Recorded | 1.25 | 2.19 | <0.001 |
| **Multimorbidity, %** | **27.7** | **37.8** | <0.001 |
| **PHRAP %** | **9.03** | **9.03** | 1 |
| **Abdominal Concern %** | **27.9** | **36.4** | <0.001 |

We first created a non-machine learning baseline prediction using a heuristic to understand if using machine learning techniques adds predictive value over more parsimonious methods. A well-performing heuristic would show that a machine learning model is an overly complex solution for little practical gain from a prediction problem. Friedman et al. found certain discharge diagnoses (**Appendix Table 2**) to be frequently associated with chief complaints of abdominal complaints in patients 65 and older.^3^ The heuristic algorithm declared any visit with any such diagnosis to be a PHRAP.

### Imputation and Feature Engineering

For the multimorbidity (*multimorb*) and total chronic conditions (*totchron*) variables, 1,240 values (< 2%) were missing within the training data set. Each missing value for one column corresponded with a missing value for the other. Total procedures (*Totproc*) and total diagnoses (*totdiag*) were missing 1,766 and 792 values within the training data set, respectively. These missing values were imputed with the mode of each column (0 for each). Principal diagnosis (*diagPrincipal*) was used for feature engineering. Specifically, *diagPrincipal* contained thousands of diagnoses, many that were irrelevant to abdominal chief complaints. We created binary variables for each of the top 78 diagnoses attributed to patients with chief complaints of abdominal pain in Friedman et al., 2021, JAGS. We also created a binary variable if any of the top 78 diagnoses were present. We scaled the continuous variable, *age*, to [0,1] using a MinMaxScaler from the Python library scikit-learn. For each patient’s age, this subtracts the minimum age in the training (2013-2017) dataset and then divides by the range of maximum age minus minimum age within the training dataset. Finally, we one-hot encoded all categorical features with more than two values and dropped the original categorical feature. One-hot encoding turns a single variable represented with three categorical values (e.g. “red”, “blue”, “green”) into three separate columns (“red”, “blue”, and “green”) which are filled by 0 for a lack of the variable and 1 for presence of the variable. For instance, a patient that had a color variable with a value of “green” at first would now have three columns: “red” (with a value of 0), “blue” (also with a value of 0), and “green” (with a value of 1).

For machine learning model training, class imbalance of non-PHRAP patients to PHRAP patients also precluded the use of accuracy as the performance metric. With a 10:1 ratio of non-PHRAP patients to PHRAP patients, a model predicting “non-PHRAP” for each patient could be accurate 90% of the time by predicting “non-PHRAP”, but fail to identify a single PHRAP patient. Instead, we chose F1 score, the harmonic mean of positive predictive value (precision) and sensitivity (recall), as our desired metric.

We used the scikit-learn Python library to perform a randomized search for hyperparameter optimization of multiple algorithms (logistic regression, random forest classifier, support vector machines, gradient boosted trees), seeking to maximize F1 score.^10^ For each set of model hyperparameters, we utilized k-fold cross-validation of our models, choosing *k* = 5. In k-fold cross-validation, the training set used to teach the machine learning model is repeatedly split into k separate mutually exclusive subsets. For each repetition, the model is trained on *k-1* subsets and validated on the *1* remaining subset. Model performance is averaged across the multiple cross-validation repetitions.^11^

Recognizing the imbalanced nature of the data could harm the predictive capacity of machine learning models, we undersampled the larger class using the Python package imbalanced-learn in order to have a lower ratio of non-PHRAP patients to PHRAP patients.^12^ The undersampling process is depicted in the supplemental materials.

After undersampling, the *k-1* subsets in each training fold the ratio of non-PHRAP to PHRAP patients was 2:1, but the non-PHRAP to PHRAP ratio remained 10:1 in the validation subset and in the final model validation of the hold-out 2018 NHAMCS data. Within the 2018 NHAMCS validation set, the “chief complaint of abdominal pain” target variable had a **10.1 : 1** ratio ((100-9.03)/9.03), and the more inclusive “abdominal concern” variable had a 1.75:1 ((100-36.4)/36.4) ratio.

#### Validation

After training on the 2013-2017 NHAMCS data, machine learning models were then asked to make predictions using patient data from 2018 NHAMCS (N=15,585) without receiving any answers. We compared the machine learning model’s predictions against the true known PHRAP/non-PHRAP labels of the 2018 NHAMCS data set to evaluate model performance, as is customary in machine learning model evaluations.

Though we only trained a model to predict chief complaints of abdominal pain, we also defined a larger, more inclusive definition of an “Abdominal Concern” to validate the model’s predictions. This more inclusive definition serves to understand the quality of the “near misses” of the PHRAP prediction algorithm. A patient could be put into this group due to their (1) reason(s) for visit to the emergency room, (2) discharge diagnosis, or (3) testing related to abdominal conditions. If a patient entered the emergency department with “Reason for Visit 1” or “Reason for Visit 2,” containing any of the complaints on the right side of **Table 1**, they were labeled as having an “Abdominal Concern.” If patients received a discharge diagnosis containing any of diagnoses associated with abdominal pain they were also added to the “Abdominal Concern” group. To develop this list, we began with the top 78 diagnoses attributed to patients with chief complaints of abdominal pain.^3^ We then eliminated diagnosis codes which did not clearly relate to the abdomen or an intra-abdominal organ. Finally, if a patient received an abdominal CT or abdominal ultrasound, they were also added to the “Abdominal Concern” group.

Though we optimized our model training for maximum F1 value, we recognize that future researchers using this model would need to trust the model’s predictions and may value higher PPV over higher sensitivity or vice-versa.

To evaluate the model, we calculated the model’s prediction probabilities ([0,1]) for each patient within the hold-out 2018 test set. Holding the prediction probabilities fixed, we could examine trade-offs in positive predictive value and sensitivity by using different decision thresholds. The default decision threshold for a scikit-learn model is 0.50, meaning if the model believes the probability of a chief complaint of abdominal pain is 0.51, it will predict a chief complaint of abdominal pain, but a probability of 0.49 would predict “not a chief complaint of abdominal pain.”

Increasing the decision threshold to 0.80, our model only predicts PHRAPs when it is extremely confident. This increases PPV but decreases sensitivity. Conversely, decreasing the decision threshold to 0.20 causes the model to predict whether a patient had a PHRAP even with relatively low confidence. This would result in a lower PPV but increased sensitivity. We plotted PPV, sensitivity, and F1 scores against various decision thresholds [0,1] and used input from clinicians to manually select a threshold that allowed for a high PPV and F1 score.

This study adheres to the TRIPOD checklist for prediction model validation (TRIPOD Checklist, 2020). It was determined to be exempt from review by the University of Pennsylvania Institutional Review Board because it uses publicly available, de-identified data.

## Results

### Data characteristics and sampling ratios

Consistent with the purposive design of the NHAMCS survey, the patient population we used for developing the PHRAP algorithm captures a broad range of ED visits, sociodemographic characteristics, and chronic health conditions. The characteristics of both the training data set and test data set populations are shown in **Table 2**, below. Coincidentally, the proportion of PHRAPs within each population is the same.

### Less-performant models

On the 2013-2017 training and validation set, the heuristic algorithm of using the presence of known diagnosis codes as a prediction for a PHRAP had a PPV of 0.24 and a sensitivity of 0.23, for an overall F1-score (the harmonic mean of PPV and sensitivity, which penalizes low values for one at the expense of the other^13^) of the PHRAPs of 0.24. The low PPV and sensitivity of this baseline indicates that without a machine learning algorithm, we would be unable to predict PHRAPs with enough accuracy to utilize for research purposes.

We found that a Light Gradient Boosting Machine implementation from Microsoft outperformed other classical machine learning methods (Random Forest, Logistic Regression, Support Vector Machine, Extreme Gradient Boosting)^14^. A simpler logistic regression provides nearly comparable results, however.

On the hold-out data set of 2018 NHAMCS patients, our final model achieved a PPV of 0.60 (95% CI of 0.56, 0.64), sensitivity of 0.29 (95% CI of 0.27, 0.32), and F1 of 0.39 for patients with PHRAPs using a decision threshold of 0.798. A decision threshold of 0.798 means that the model must have 79.8% confidence that the patient has a PHRAP to predict a PHRAP.

Of a holdout test set of 15585 patients, our model had 1,000 false negatives (6.4% of total) and 271 false positives (1.7% of total). We found that 99.3% of false positives (patients predicted to have PHRAP but not actually entering the emergency department with a PHRAP) presented to the emergency department with abdominal concerns as defined in Table 1 (Figure 1). Of 271 false positives, 108 (39.9%) patients exhibit abdominal pain as a secondary complaint for their visit to the emergency department, 159 (58.7%) received an abdominal discharge diagnosis, and 264 (97.4%) received an abdominal CT scan or ultrasound. Only 2 patients (0.7%) from the original false positive group entered the ED without any abdominal concern. These two false positives were reviewed by a practicing emergency physician (ABF) and confirmed to be non-instances of abdominal pain.

**Figure 1.**
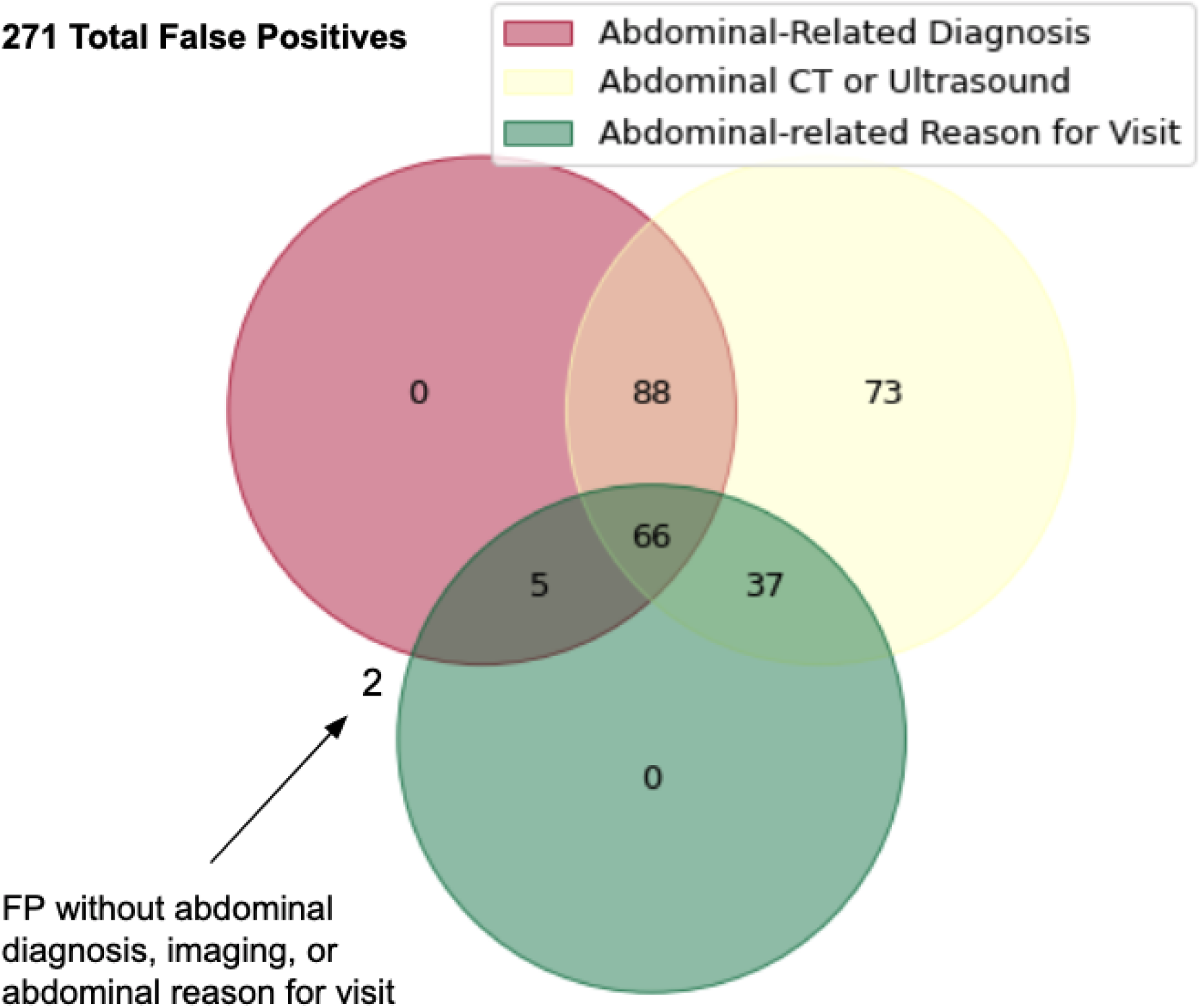
Venn Diagram of False Positive PHRAP Predictions.

Figure 2 compares positive predictive value to sensitivity across different thresholds for the model’s predictions of PHRAPs (blue) and its predictions of PHRAPs compared to the broader classification of “abdominal concerns” (orange). The predictions don’t change between the two lines, but the blue line compares these predictions to true PHRAPs while the orange line compares the model’s predictions to the label of “abdominal concern.” The orange “abdominal concern” line shows substantially better performance than the blue “PHRAP” line, demonstrating that when the model “misses” in predicting a PHRAP when in reality there is none, the model is making these predictions in cases where the patient still has abdominal concerns. This may be reassuring for future researchers using this model in claims data who want to define a patient population entering the emergency department with abdominal pain.

**Figure 2.**
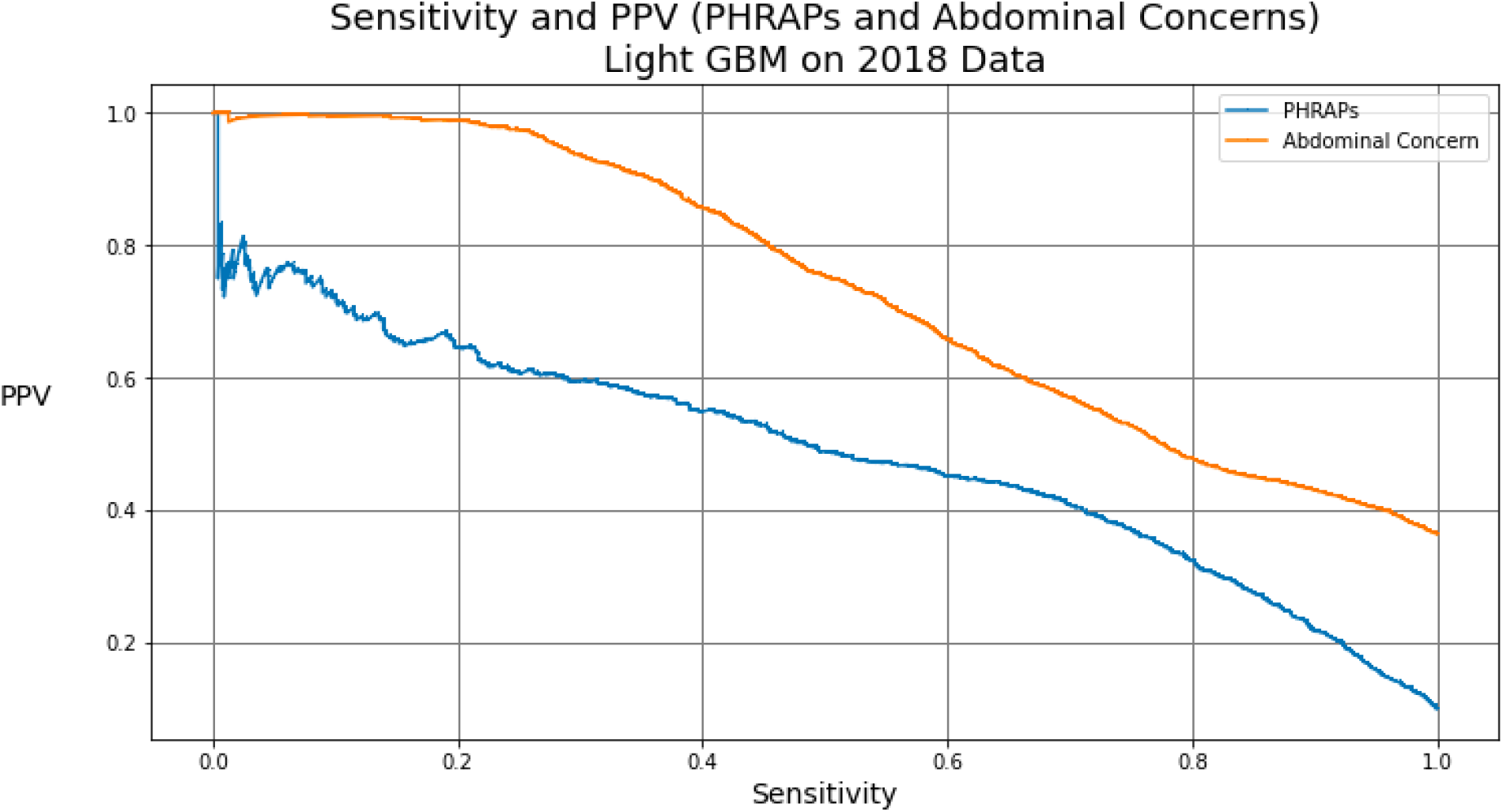
Precision and recall curves of best model for PHRAP and Abdominal Concern.

**Figure 3.**
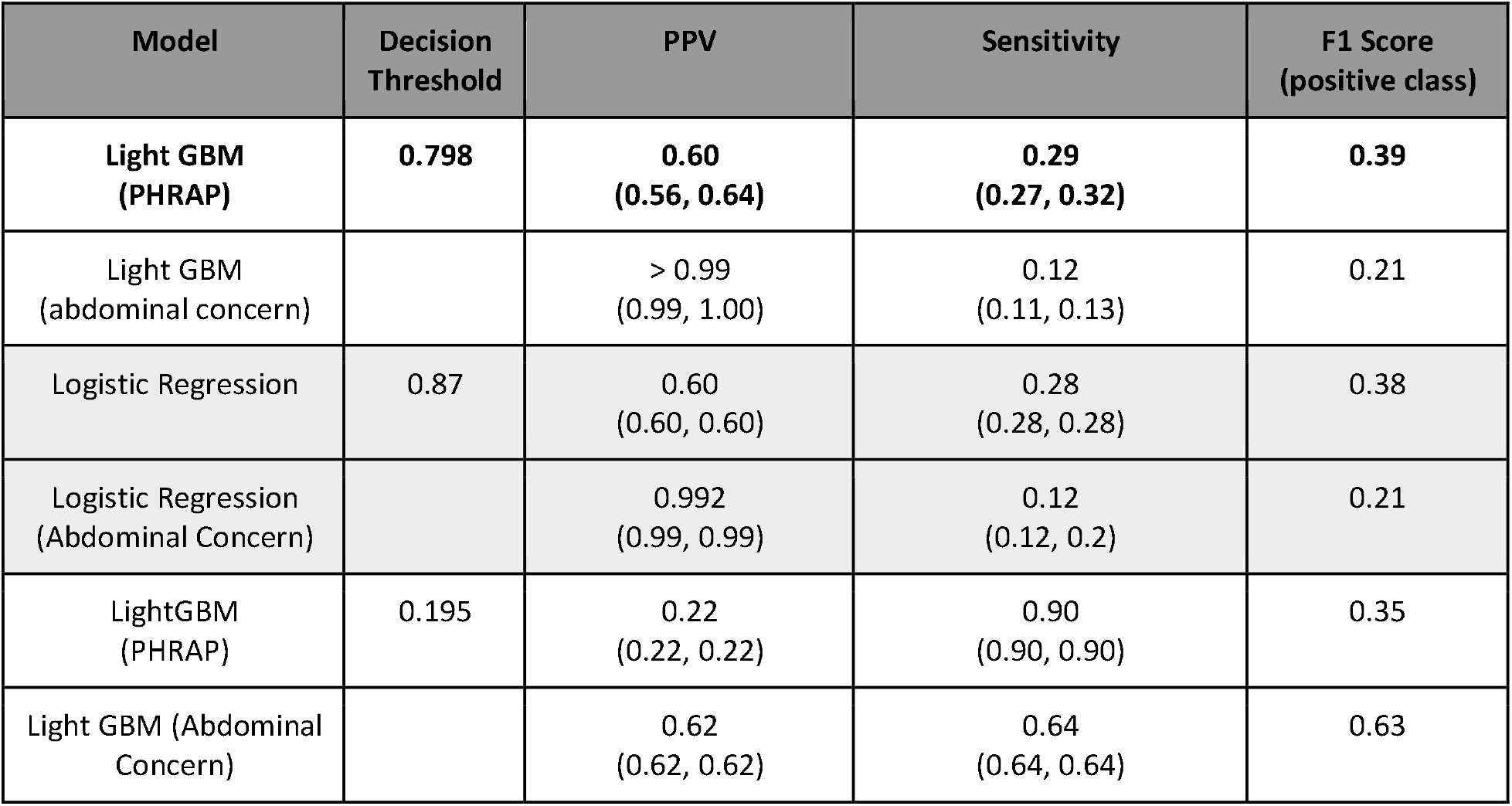
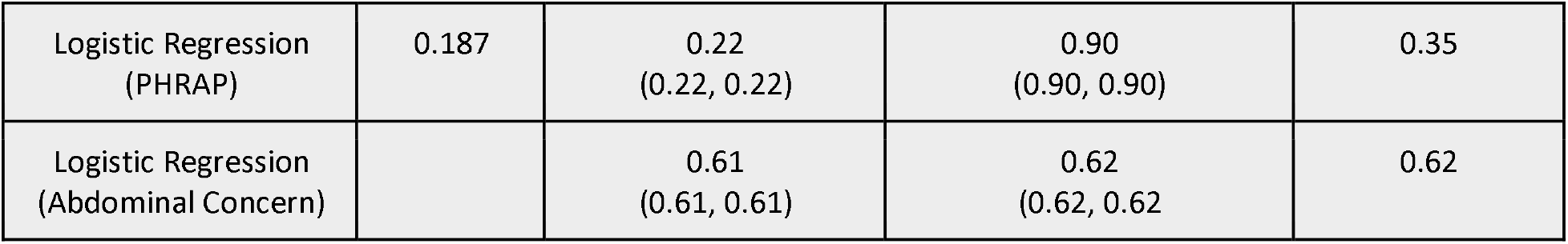
Performance Measures for best model.

Table 3 depicts the results of the best predictive model as well as a comparison to a simpler logistic regression model. For both models, we prefer a decision threshold that gives a high PPV at the cost of sensitivity. We also include a prediction decision threshold that supports higher sensitivity at the cost of PPV.

## Discussion

We developed a machine learning model to predict chief complaints of abdominal pain within NHAMCS data from 2013-2017 using only variables commonly available in claims data. We validated our model on a hold-out set of unseen test data from 2018. The inherent variability of clinical data due complex biology, varied communication styles, and variegated clinical judgment makes high performance of both PPV and sensitivity difficult, so we offer two separate models, one high PPV and one with high sensitivity. We show the usefulness of the PHRAP model in defining a PHRAP patient population through its relatively high PPV (0.6) and in examination of the false positive errors—99% of which are patients with complaints, diagnoses, or testing consistent with abdominal concerns. We find that a simpler logistic regression model performs nearly as well as the best model, a gradient-boosted machine. This is consistent with previous research that machine learning approaches do not always perform better than logistic regression models in clinical prediction tasks.^15^

Older patients presenting to an ED with a chief complaint of abdominal pain is a high mortality, high morbidity condition. While national care patterns for this high-risk population are known, the impact of those care decisions in the ED on outcomes are not, because post-discharge outcomes data such as 30-day mortality is not available in ED-centric databases which include a chief complaint. We developed a method of defining a population who presented to the ED with a potentially high-risk abdominal presentation.

The model’s high PPV and “near miss” detection of abdominal concerns help researchers define a patient population within claims data that has potentially high risk abdominal presentations (PHRAPs) when entering the emergency department. Because a simpler logistic regression model performs nearly as well as a gradient boosted machine, we support using the logistic regression model and provide the model coefficients in an appendix. By defining this patient population, future researchers can better study claims data to more accurately quantify the risks of patients entering the emergency department with abdominal complaints, define metrics for treatment success, and create care pathways for these high-risk cases.

Beyond the domain of abdominal pain, we see promise in the use of predictive machine learning models to define other populations in claims data and thereby link clinical outcomes to clinical testing and patient complaints upon entering the emergency department.

Because no curated data set linking chief complaints of abdominal pain and associated testing and imaging to outcomes exists, our prediction model attempts to bridge the gap. Though we have high confidence in this model to segment an appropriate patient population within claims data due to our model’s high PPV and the high prevalence of abdominal concerns within the false positive predictions, the predictions must be viewed as a tool for research of populations rather than any kind of clinical diagnostic tool for patient treatment.

## Data Availability

Source data is publicly available: https://www.cdc.gov/nchs/ahcd/datasets_documentation_related.htm
All data produced in the present study are available upon reasonable request to the authors.

